# A simple method to quantify country-specific effects of COVID-19 containment measures

**DOI:** 10.1101/2020.04.07.20057075

**Authors:** Morten Gram Pedersen, Matteo Meneghini

## Abstract

Most of the world is currently fighting to limit the impact of the COVID-19 pandemic. Italy, the Western country with most COVID-19 related deaths, was the first to implement drastic containment measures in early March, 2020. Since then most other European countries, the USA, Canada and Australia, have implemented similar restrictions, ranging from school closures, banning of recreational activities and large events, to complete lockdown. Such limitations, and softer promotion of social distancing, may be more effective in one society than in another due to cultural or political differences. It is therefore important to evaluate the effectiveness of these initiatives by analyzing country-specific COVID-19 data. We propose to model COVID-19 dynamics with a SIQR (susceptible – infectious – quarantined – recovered) model, since confirmed positive cases are isolated and do not transmit the disease. We provide an explicit formula that is easily implemented and permits us to fit official COVID-19 data in a series of Western countries. We found excellent agreement with data-driven estimation of the day-of-change in disease dynamics and the dates when official interventions were introduced. Our analysis predicts that for most countries only the more drastic restrictions have reduced virus spreading. Further, we predict that the number of unidentified COVID-19-positive individuals at the beginning of the epidemic is ∼10 times the number of confirmed cases. Our results provide important insight for future planning of non-pharmacological interventions aiming to contain spreading of COVID-19 and similar diseases.

## Introduction

The COVID-19 disease due to the SARS-nCov2 coronavirus is spreading rapidly across the globe since its outbreak in China, and was declared a pandemic by the WHO on March 11, 2020. After the first severe patient was brought to the hospital of Codogno, Italy on February 20, 2020, and subsequently tested positive for SARS-nCov2, a rapidly increasing number of patients have been identified, initially in Northern Italy and later in the rest of the country and Europe. Italy is the most affected Western country, with more than 130.000 confirmed cases, a death toll of ∼16.000, and the first to implement drastic measures in an attempt to contain the disease. Since then, most other European countries, the USA and Canada, have implemented similar restrictions, ranging from school closures, banning of recreational activities and large events, to complete lockdown. In order to evaluate the effectiveness of these initiatives, it is necessary to analyze COVID-19 data carefully to investigate if and how the limitations in societal activities affect the disease dynamics.

It is not obvious that specific interventions have the same effects in all countries. For example, school closures may have secondary effects concerning “smart working” of the parents staying at home with the children. However, the possibility to work from home depends on the type of job, so that school closures may have a bigger effect in countries with more tertiary sector jobs compared to, for example, factories. Further, there are social aspects that come into play, such as the possibility of grandparents to care for the children when schools close, which makes it possible for parents to continue working, but also expose the older citizens, who are the most vulnerable to COVID-19, to the disease. Naturally, also the relative number of children in the society play a role. Another important aspect to consider concerns national and regional cultural and psychological differences for example regarding respect versus authorities. If the general population does not trust the communications coming from health and political authorities, or if these communications are confusing and contradictory, more stringent bans can be necessary in order to contain the disease, since softer suggestions may not be followed.

Simple mathematical models of infectious diseases are useful for providing insight into disease dynamics, and compared to more complex models, can be fitted to data with a minimum number of assumptions on model parameters. This fact is particularly important when analyzing data from a series of countries without assuming for example that specific interventions have the same effect in all countries. However, even simple models should respect that nature of the data. There is thus a compromise between using a parsimonious model but sufficiently complex to be based on correct underlying assumptions.

In our setting, to fit the data on identified SARS-nCov2 positive cases, a SIQR [1] is appropriate. In this model, infected individuals may be isolated, entering the “quarantined” subpopulation Q, so that these individuals no longer transmit the disease. Since positive cases are put in isolation (in hospitals or at home) immediately, the revealed data of active cases thus corresponds to the number of individuals in state Q. It would not be correct to confront the infectious subpopulation of a SIR/SEIR (susceptible – infectious – [exposed] – recovered) model with the recorded data, since it is unknown how many infectious but undetected individuals are circulating in the population. As will be clear in the following, besides being conceptually clearer, the explicit quantification of the quarantined individuals will allow us to provide estimates of the infectious cases at the very beginning of the epidemic outbreak as well as of the current number of undetected infectious individuals.

We show that our relatively simple approach is able to capture and quantify the effects of restrictions in a series of Western countries. In contrast to previous modelling approaches [2], we do not assume that interventions have effect on a particular date, or have the same effect in all countries. Our model can thus estimate for example if the effect of an interventions is delayed with respect to the date of implementation, or if the disease dynamics changed earlier, for example because of public awareness of the necessity of restricting social interactions, thus anticipating formal bans.

## Results

To parameterize the SIQR model, we fitted analytically derived expressions for *Q* + *R* (see Methods) to the data of total COVID-19 cases in seven different European countries, British Columbia (Canada), New South Wales (Australia) and New York City (USA), from the first date when the number of confirmed cases of the single regions passed above 50 through April 2, 2020. (Fig. 1). The underlying model was assumed to have a piecewise-constant transmission rate *β*, taking one value before a given date and another value after that date (date-of-change in the following). This assumption on *β* translates into a piecewise constant growth rate for the number of infectious individuals *I* (see Methods). Table 1 shows estimated values for the four free, identifiable parameters of the model: date-of-change (*T*_*c*_), growth rates before 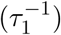 and after 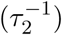 the date-of-change, and the combined parameter *ηI*_0_ of quarantine rate (*η*) times the initial value of the number of non-identified positive cases (*I*_0_).

**Table 1:**
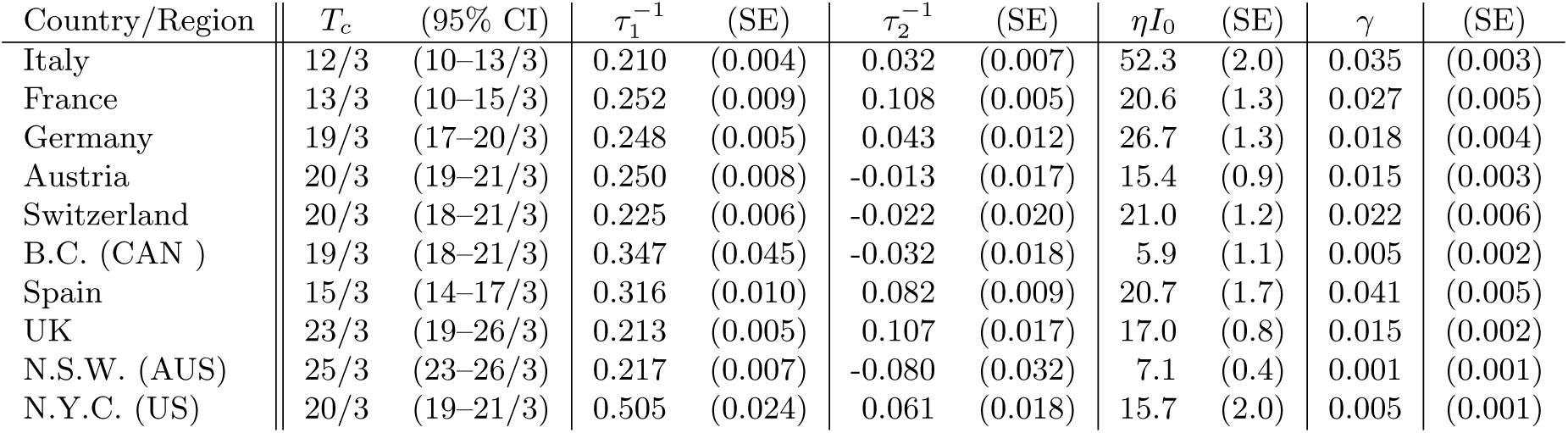
Estimated model parameters for the different countries and regions: Date-of-change (*T*_*c*_) and dates falling within the 95% confidence interval (CI), growth rates before 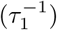 and after 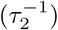 the date-of-change with standard errors (SE), the quarantine rate times the initial number of infectious cases (*ηI*_0_) with SE, and the combined death/recovery rate (*γ*) with SE.

| Country/Region | $T_c$ | (95% CI) | $\tau_1^{-1}$ | (SE) | $\tau_2^{-1}$ | (SE) | $\eta I_0$ | (SE) | $\gamma$ | (SE) |
| --- | --- | --- | --- | --- | --- | --- | --- | --- | --- | --- |
| Italy | 12/3 | (10–13/3) | 0.210 | (0.004) | 0.032 | (0.007) | 52.3 | (2.0) | 0.035 | (0.003) |
| France | 13/3 | (10–15/3) | 0.252 | (0.009) | 0.108 | (0.005) | 20.6 | (1.3) | 0.027 | (0.005) |
| Germany | 19/3 | (17–20/3) | 0.248 | (0.005) | 0.043 | (0.012) | 26.7 | (1.3) | 0.018 | (0.004) |
| Austria | 20/3 | (19–21/3) | 0.250 | (0.008) | -0.013 | (0.017) | 15.4 | (0.9) | 0.015 | (0.003) |
| Switzerland | 20/3 | (18–21/3) | 0.225 | (0.006) | -0.022 | (0.020) | 21.0 | (1.2) | 0.022 | (0.006) |
| B.C. (CAN ) | 19/3 | (18–21/3) | 0.347 | (0.045) | -0.032 | (0.018) | 5.9 | (1.1) | 0.005 | (0.002) |
| Spain | 15/3 | (14–17/3) | 0.316 | (0.010) | 0.082 | (0.009) | 20.7 | (1.7) | 0.041 | (0.005) |
| UK | 23/3 | (19–26/3) | 0.213 | (0.005) | 0.107 | (0.017) | 17.0 | (0.8) | 0.015 | (0.002) |
| N.S.W. (AUS) | 25/3 | (23–26/3) | 0.217 | (0.007) | -0.080 | (0.032) | 7.1 | (0.4) | 0.001 | (0.001) |
| N.Y.C. (US) | 20/3 | (19–21/3) | 0.505 | (0.024) | 0.061 | (0.018) | 15.7 | (2.0) | 0.005 | (0.001) |

**Figure 1:**
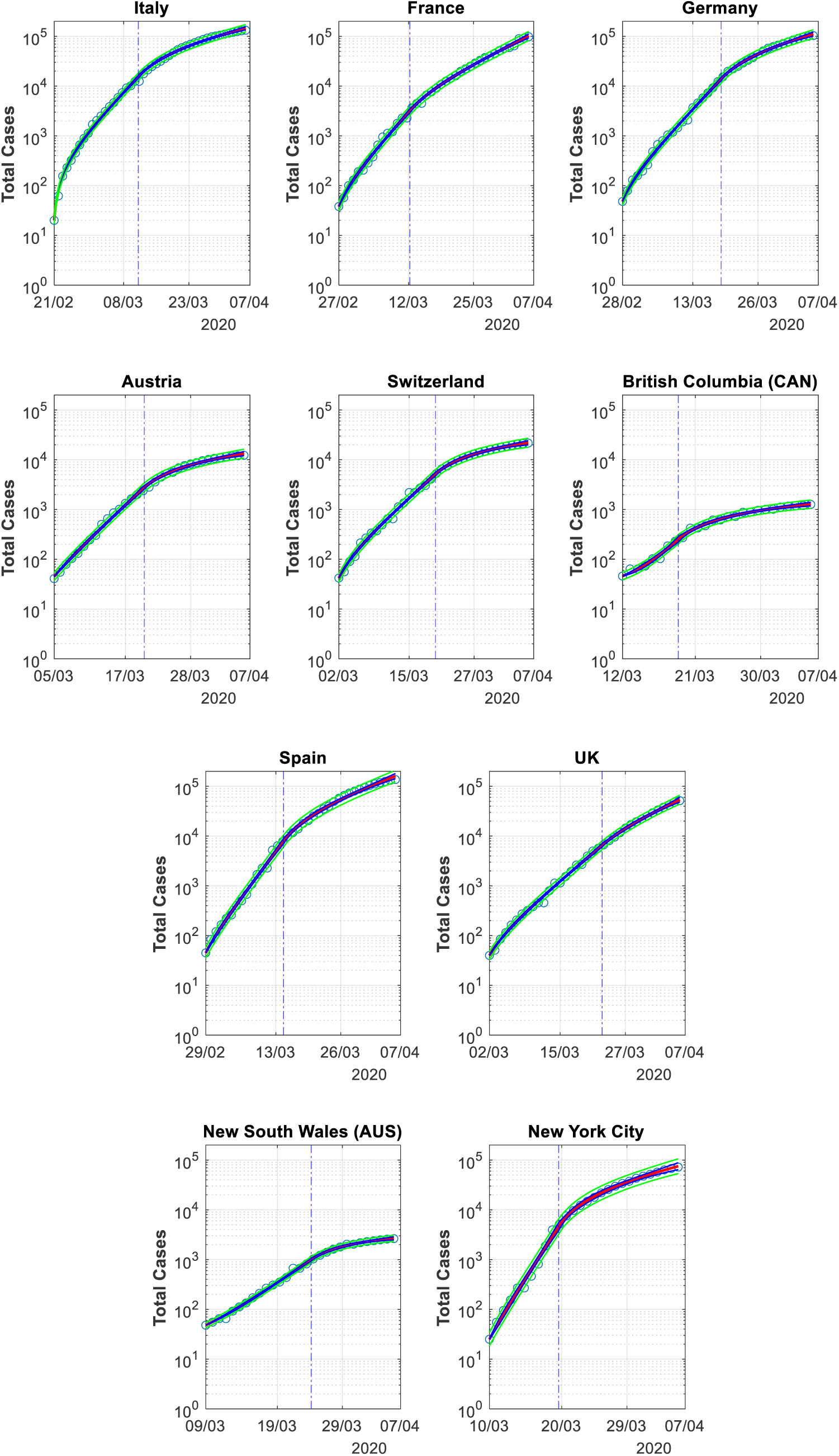
The total number of COVID-19 positive cases (circles) and the best fits (red curve) with piecewise constant growth rates 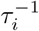, *i* = 1, 2. The date-of-change is indicated by the vertical dashed line, 95% confidence bands are given in blue and 95% prediction bands in green. Data from https://github.com/pcm-dpc/COVID-19 and https://github.com/CSSEGISandData/COVID-19.

Most countries and regions initially had similar growth rates of 0.2–0.3/day (Fig. 2), corresponding to a doubling time of ∼2.5–3.5 days. However, growth in New York City (N.Y.C.) was much faster with a doubling time of ∼1.4 days. After the date-of-change all regions lowered the growth rate significantly (*p <* 0.001). We estimated that the growth rate in Austria, Switzerland, and British Columbia (B.C.) is practically zero, i.e., the basic reproduction number *R*_0_ ≈ 1, whereas New South Wales (N.S.W.) have statistically significant negative growth rate (*p* = 0.02). Italy, Germany, and N.Y.C. have growth rates of ∼0.03–0.06/day, i.e. doubling times of 11–22 days. In a more detailed analysis with two dates-of-change (not shown here) we have estimated that Italy more recently have lowered the growth rate to ∼0/day following closure of job activities deemed not strictly necessary. France, Spain and the UK still have growth rates of 0.08– 0.11/day (doubling times 6.5-8.5 days).

**Figure 2:**
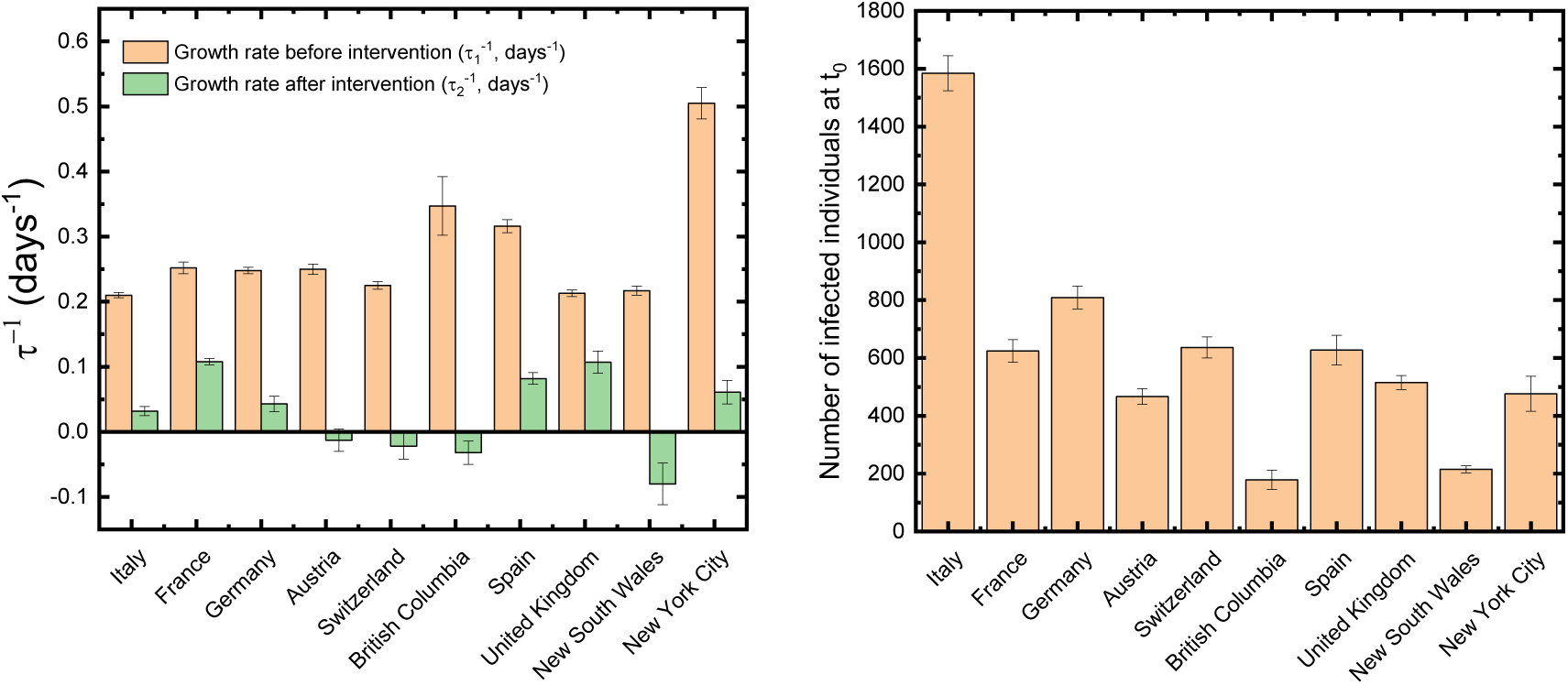
Overview of the estimated growth rates before and after the date-of-change (left), and the number of infectious individuals (*I*_0_, right) at the beginning of the outbreak.

Table 2 shows when major restrictions were introduced in the different countries. We notice an excellent agreement between data-driven estimation of *T*_*c*_ and the dates of major public interventions, in particular school closings or partial or complete lockdown. However, there are interesting differences between the countries. For example, in Italy disease dynamics changed only after complete lockdown whereas in New South Wales (Australia), COVID-19 was contained in response to suggested, not ordered, absence from school. These differences point to important cultural, political or societal differences between the countries. We will return to this crucial point in the Discussion.

**Table 2:** Confrontation between estimated date-of-change (*T*_*c*_; from Table 1) and major containment measures. Asterisks indicate local restrictions in part of the country.

| Country/Region | $T_c$ | (95% CI) | School closures | Lockdown |
| --- | --- | --- | --- | --- |
| Italy | 12/3 | (10–13/3) | 22/2*, 5/3 | 9/3*, 11/3 |
| France | 13/3 | (10–15/3) | 13/3*, 16/3 | 15/3, 17/3 |
| Germany | 19/3 | (17–20/3) | 16/3*, 18/3 | 16/3, 19/3 |
| Austria | 20/3 | (19–21/3) | 17/3 | 17/3 |
| Switzerland | 20/3 | (18–21/3) | 16/3 | 17/3 |
| B.C. (CAN ) | 19/3 | (18–21/3) | 17/3 | 20/3 |
| Spain | 15/3 | (14–17/3) | 11/3*, 16/3 | 14/3 |
| UK | 23/3 | (19–26/3) | 20/3 | 21/3 |
| N.S.W. (AUS) | 25/3 | (23–26/3) | 23/3 (recommended) | 31/3 |
| N.Y.C. (US) | 20/3 | (19–21/3) | 16/3 | 20/3 |

Our model has only two free parameters to be determined from other data. The estimated values for the cure rate of non-identified cases, *α* = 0.067/day, and the rate of isolation, *η* = 0.033/day, (see Methods) yield *β* = *τ*^*−*1^ + 0.1 and basic reproduction number *R*_0_ = *τ*^*−*1^*/*0.1 + 1, which falls between 3.1 and 4.5 for all regions except N.Y.C. (*R*_0_ = 6.1) for the initial period. These values are in line with previous estimates of *R*_0_ lying between 2 and 4 [3–7]. After the restrictions were introduced, *R*_0_ dropped to 0.2 – 2.1.

Further, we obtain an estimate of the number of infectious individual at the moment of the outbreak in the different countries and regions (Fig. 2). Assuming that *η* is identical for all countries we estimate that there are large differences in this number. For Italy, the first European country hit by COVID-19, we estimate that as many as ∼1500 individuals were infectious but unidentified at the moment of the outbreak, possibly because the outbreak of the disease was considered an unlikely event in the country, which therefore did not test patients with milder COVID-19 symptoms. In other European countries (Spain, France, Germany, Switzerland), which saw the epidemic arriving a few days later than Italy, the estimated initial number of infectious individuals is 600–800. In contrast, for B.C. in Canada and N.S.W. in Australia, which had more time to prepare for COVID-19, this number was only ∼200 when the number of confirmed cases reached 50.

We simulated the dynamics of the SIQR model (Fig. 3) and found that the data of the different countries was very well fitted when assuming piecewise-constant time-variance in *β*. From Eq. 3 and Eq. 5 (see Methods) it can be shown analytically that following the transient and during the initial exponential growth, 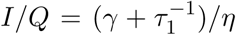, where the combined recovery/death rate *γ* can be estimated from data (see Methods). In other words, the relation between *Q* and *I* follows directly from the data up to the factor *η*. With the value of *η* = 0.033/day that we use here, we obtain that, e.g., for Italy the number of hidden infectious individuals is ∼7 times the number of positive cases, whereas in N.Y.C. this number is as high as ∼15. In the other countries the value is 6-10. These *I*-to-*Q*-ratios are slightly higher than the result of a recent study on the number of unidentified infectious individual during the epidemic in China [8], but in line with a study on the European countries [2].

**Figure 3:**
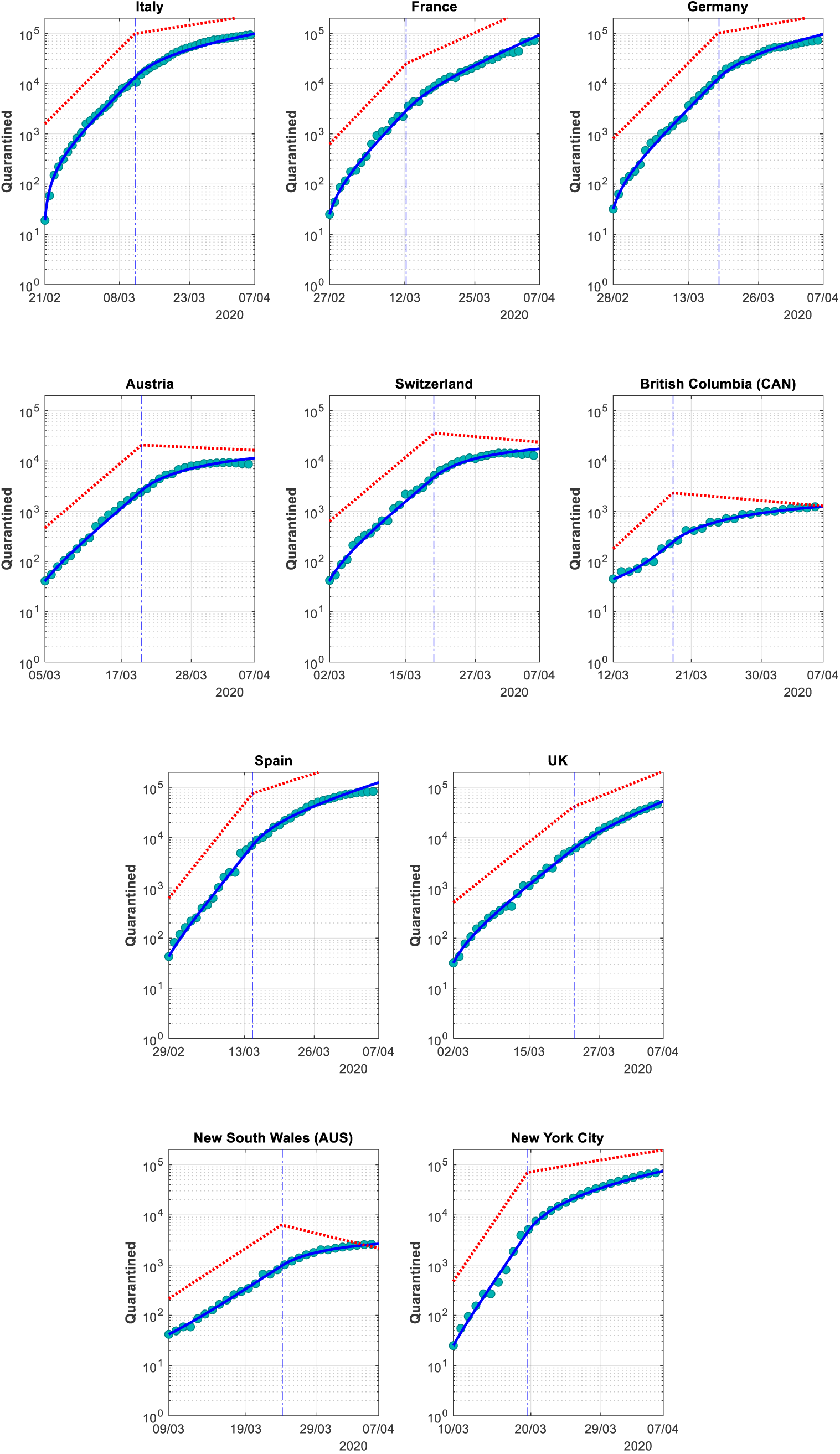
Simulation of the SIQR model with parameters as given in Table 1. The number of active cases (calculated as confirmed minus recovered and deceased cases) are shown by the dots. The blue curves show the simulated number of quarantined cases *Q*. The red, dashed curves indicate the simulated number of infectious but hidden cases, *I*.

## Discussion and conclusions

We have presented a mathematically simple framework for the study of the effects of public interventions in specific countries, regions or cities based on a SIQR model. Remarkably, we show that for all analyzed countries our model can effectively estimate the date in which the measures started being effective, thus allowing to assess the efficacy of the interventions put in place by the different governments and the response of the populations.

Interestingly the regions that have lowered the growth rate to zero have imposed relatively mild restrictions. In New South Wales schools are not closed, but parents were invited to keep their children at home on March 23, 2020. This date coincides almost perfectly with our estimated date-of-change, suggesting that this invitation has been followed, and has had immediate effect. Indirect effects likely contribute: keeping children at home often means that their parent will work from home, and in addition, the increased awareness of the situation following such an invitation may have wider, general effects, as might be expected to have occurred in N.S.W. Similarly, B.C. has not imposed lockdown, although schools were closed on March 18, 2020 (in agreement with our estimated date-of-change) and restaurants are only allowed to sell take-away food.

Switzerland, in spite of its closeness to Italy, has managed to contain COVID-19 without a strict lockdown as seen in Italy. In Switzerland, schools closed on March 16, 2020, and a partial lockdown involving closure of bars, shops and other gathering places, but leaving open certain essentials, such as grocery stores, pharmacies, (a reduced) public transport and the postal service, was imposed from March 17, 2020. We estimate that these interventions had effect on the COVID-19 transmission rate within a few days.

Our analysis of Germany and Austria in many aspects mimic Switzerland, except that for Germany the growth rate is still positive (*R*_0_ > 1). Our estimated date-of-change coincides with the dates of school closures and partial lockdown. Most German federal states closed schools from March 16, 2020 followed by the remaining country from March 18, 2020 – one day before our estimated date of change. The German government imposed partial lockdown from March 17, closing all non-essential shops, and imposing limitation restaurants and banning leisure activities, which was followed by further restrictions in the following days. On March 16 the government of Austria prohibited gatherings of groups of more than five people in public, and all schools were closed from March 17. On this latter date, the closure of restaurants was also ordered. People were recommended to leave home only for purchasing necessary goods (medications, groceries), for assisting other people, or for important professional activities. The date-of-change estimated by our model (March 20) corresponds nicely to the dates of these restrictions, demonstrating their effectiveness in limiting the growth rate of the epidemics.

For Italy we saw no effect of school closures. However, the worst hit Northern regions closed schools immediately after the first cases were registered on February 20-21, 2020. These dates coincides with the beginning of the carnival school holidays. Hence, the date of school closure may not show up in the data since schools were shut anyway. The fact that the initial estimated growth rate in Italy is slightly slower than in e.g. France and Spain may reflect that schools were closed from the beginning of the epidemic in Italy. For Italy, we estimated that the growth rate changed on 12/3, coinciding with the complete lockdown that was introduced March 9–11, 2020.

France closed schools between March 13–16, 2020, first in the worst hit areas and then nationally. Our estimated date-of-change, March 13, suggests that school closure in the worst hit regions had immediate effect. March 17, the date of further lockdown, did not show up in our data analysis but these additional restrictions may nonetheless contribute to the overall lowering of the growth rate. In Spain and UK, the estimated dates-of-change coincide with the lockdowns introduced March 14, 2020, respectively March 21, 2020, which were accompanied by school closures within a few days.

In spite of the lockdowns in France, Spain and the UK, the estimated growth rates are still high. There might be important social, political and communicative differences between these countries and e.g. Switzerland, B.C., N.S.W. and Germany, which can explain how more stringent measures have had less effect.

On a more methodological side, our analysis clarifies that the early decrease in slope seen in the log-transformed data (Fig. 1) is not due to early containment measures, but a direct consequence of the model structure. Further, our model approach, which notably has only two free parameters to estimate, allows us to estimate that the number of infectious individuals *I* is approximately 10 times larger than the number of isolated patients *Q* (Fig. 3). This fact follows to a large extent directly from the data, as explained above.

We were also able to estimate that there were ∼1500 of infectious but undetected infectious individuals in Italy at the time of the outbreak around February 21, 2020. Only when a patient with severe symptoms was hospitalized and tested for SARS-nCov2, and the first infected person died from COVID-19 on the following day, wide testing and isolation efforts started, which revealed a large basin of overlooked cases. In contrast, we found a much lower number of hidden cases for New South Wales (Australia) and British Columbia (Canada) at the time of the outbreak, likely because these countries had more time to prepare for the epidemic and starting testing for COVID-19 early.

In summary, our parsimonious modelling approach permits extracting important information on the efficacy of COVID-19 containment measures in individual countries, regions and cities. These results can then be analyzed further to understand why some measures are working in one societal setting but is less effective in another. Such insight would help future planning of non-pharmacological interventions aiming to contain spreading of COVID-19 and similar diseases.

## Methods

We use a SIQR model [1] to describe COVID-19 dynamics in Italy. The model equations are

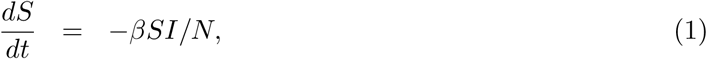

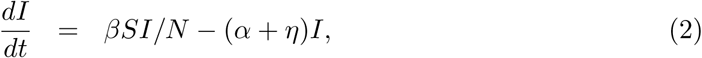

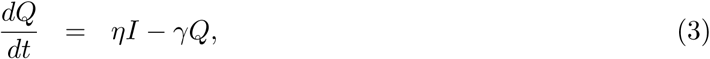

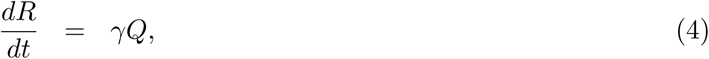

where *R* models SARS-nCov2-positive, isolated individuals that recover or die from the disease. We do not explicitly model the number of recovered or deceased non-identified COVID patients, but only the rate *α* with which these patient become non-infectious. Further, *N* is the total number of individuals in the population, assumed constant since we are studying the early phase of the epidemic.

Since there is evidence that the SARS-nCov2 virus can be transmitted in the absence of symptoms [9, 10], we do not include an explicit exposed-but-noninfectious (E) state, i.e., we do not consider a SEIQR model [11]. Further, from the Diamond Princess cruiseship, it has been found that ∼50% of SARS-nCoV2-positive individuals do not develop symptoms [12]. Similarly, complete testing in the village of Vo’ Euganeo showed that 50-75% of infected individuals were asymptomatic [13]. We assume that such positive but asymptotic individuals can transmit the disease, i.e., the *I* state includes both individuals that will not develop symptoms, cases that did not develop symptoms yet, and symptotic patient that still have not been tested positive and isolated.

So far a relatively small fraction of the Italian population has been found positive for COVID-19. Thus, we are still in the early phase of the epidemic where *S* ≈ *N*, and as well known in the case of constant parameters, the number of infected individuals follows exponential growth

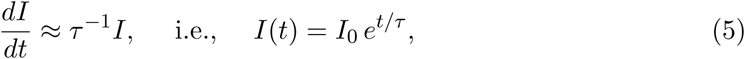

where *I*_0_ is the number of infectious individuals at the beginning of the outbreak *t* = 0 and *τ*^*−*1^ = *β* − (*α* + *η*) is the growth rate.

As mentioned, the number of individuals that have been found SARS-nCov2 positive and put in isolation does not correspond to *I* but to *Q* + *R*. From Eq. 5 it follows that *Q* + *R* follows

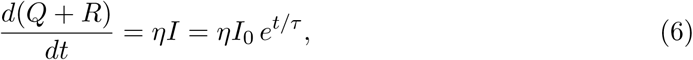

which yields

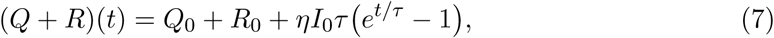

where *Q*_0_ and *R*_0_ are initial values of *Q* and *R*, respectively. Assuming that the growth rate changes from 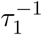 to 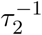 on the date-of-change *T*_*c*_, we obtain

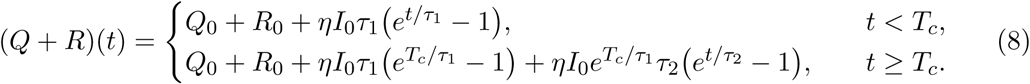

Using the nls function in R [14], we fitted Eq. 8 with free parameters *T*_*c*_, 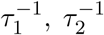, and (*ηI*_0_), to the log-transformed COVID-19 data for each country or region, from the first date when the number of infected cases passed above 50 until April 2, 2020. Confidence and prediction bands were calculated in Origin.

The recovery/death rate *γ* was obtained, for each country, by averaging the day-to-day differences in the number of recovered/deceased cases (Δ*R*_*i*_ = *R*_*i*_ − *R*_*i−*1_) divided by the number of active cases on the corresponding day (*Q*_*i−*1_), i.e., *γ* = avg(Δ*R*_*i*_*/Q*_*i−*1_). The values obtained are reported in Table 1.

To identify the other individual parameters we use previous findings. It has been estimated that the average incubation time is ∼5 days [3, 15] and the duration of the milder cases of disease it 5-10 days [9]. We assume an average time of duration from infection to recovery or death of non-isolated cases of 10 days, corresponding to a rate of 0.1/day. If a fraction *δ* of infectious individuals is tested positive and put in isolation, we obtain *α* = (1 − *δ*)× 0.1/day.

Similarly *η* related to the time until patients are tested positive and isolated, but also to the fraction of all infectious individuals that are tested positive. These are mostly symptomatic patients, which we assume are isolated 5 days after the incubation time is over and first symptoms appear, i.e., after ∼10 days. Letting *δ* denote the fraction of infectious individual entering *Q*, we obtain *η* = *δ* × 0.1/day.

Since ∼50–75% of the population is asymptomatic [12, 13], and some milder cases may also go unnoticed and not end in isolation, we assume that *δ* = 1*/*3 of infectious individuals are tested after an average of 10 days. We thus obtain *η* = 0.033/days and *α* = 0.067/days.

## Data Availability

Public available data only. URLs in the paper.

## Authors contributions

MGP and MM conceived research and discussed all results. MGP developed the theoretical framework and performed parameter estimation. MM developed the simulation framework and compared real data to model results. MGP wrote the paper. MM commented drafts and approved the final version of the paper.

## References

[1] Hethcote H, Zhien M, Shengbing L. Effects of quarantine in six endemic models for infectious diseases. Mathematical biosciences. 2002;180(1-2):141–160. doi:10.1016/S0025-5564(02)00111-6

[2] Flaxman S, Mishra S, Gandy A, et al. Estimating the number of infections and the impact of nonpharmaceutical interventions on COVID-19 in 11 European countries. Imperial College COVID-19 Response Team. Mar 30, 2020. doi: 10.25561/77731.

[3] Li Q, Guan X, Wu P, Wang X, Zhou L, Tong Y, et al. Early Transmission Dynamics in Wuhan, China, of Novel Coronavirus-Infected Pneumonia. N Engl J Med. 2020 Jan;. doi:10.1056/NEJMoa2001316

[4] Zhou T, Liu Q, Yang Z, Liao J, Yang K, Bai W, et al. Preliminary prediction of the basic reproduction number of the Wuhan novel coronavirus 2019-nCoV. J Evid Based Med. 2020 Feb;13(1):3–7. doi:10.1111/jebm.12376

[5] Zhao S, Lin Q, Ran J, Musa SS, Yang G, Wang W, et al. Preliminary estimation of the basic reproduction number of novel coronavirus (2019-nCoV) in China, from 2019 to 2020: A data-driven analysis in the early phase of the outbreak. International Journal of Infectious Diseases. 2020;92:214–217. doi:10.1016/j.ijid.2020.01.050.

[6] Liu Y, Gayle AA, Wilder-Smith A, Rocklöv J. The reproductive number of COVID-19 is higher compared to SARS coronavirus. Journal of travel medicine. 2020;. doi:10.1093/jtm/taaa021

[7] Lai A, Bergna A, Acciarri C, Galli M, Zehender G. Early phylogenetic estimate of the effective reproduction number of SARS-CoV-2. Journal of medical virology. 2020;. doi:10.1002/jmv.25723

[8] Li R, Pei S, Chen B, Song Y, Zhang T, Yang W, et al. Substantial undocumented infection facilitates the rapid dissemination of novel coronavirus (SARS-CoV2). Science. 2020 Mar;. doi:10.1126/science.abb3221

[9] Bai Y, Yao L, Wei T, Tian F, Jin DY, Chen L, et al. Presumed asymptomatic carrier transmission of COVID-19. Jama. 2020;. doi:10.1001/jama.2020.2565

[10] Rothe C, Schunk M, Sothmann P, Bretzel G, Froeschl G, Wallrauch C, et al. Transmission of 2019-nCoV infection from an asymptomatic contact in Germany. N Engl J Med. 2020;382:970–971. doi:10.1056/NEJMc2001468

[11] Jumpen W, Wiwatanapataphee B, Wu Y, Tang I. A SEIQR model for pandemic influenza and its parameter identification. International Journal of Pure and Applied Mathematics. 2009;52(2):247–265.

[12] National Institute of Infectious Diseases J. Field Briefing: Diamond Princess COVID-19 Cases, 20 Feb Update;. Available from: https://www.niid.go.jp/niid/en/2019-ncov-e/9417-covid-dp-fe-02.html.

[13] Day M. Covid-19: identifying and isolating asymptomatic people helped eliminate virus in Italian village. BMJ. 2020 Mar;368:m1165. doi:10.1136/bmj.m1165

[14] R Core Team. R: A Language and Environment for Statistical Computing. Vienna, Austria; 2016. Available from: https://www.R-project.org/.

[15] Lauer SA, Grantz KH, Bi Q, Jones FK, Zheng Q, Meredith HR, et al. The Incubation Period of Coronavirus Disease 2019 (COVID-19) From Publicly Reported Confirmed Cases: Estimation and Application. Ann Intern Med. 2020 Mar;. doi:10.7326/M20-0504

